# Process evaluation of the INTENSE study aimed at improving adherence to oral diabetes and/or hypertension medication in people with type 2 diabetes

**DOI:** 10.64898/2026.10.28.26366708

**Authors:** Marlous Langendoen-Gort, Debi Bhattacharya, Hiyam Al-Jabr, Maartje de Wit, Jacqueline G Hugtenburg, Andrew Farmer, Petra JM Elders, Femke Rutters

**Affiliations:** Amsterdam UMC, location Vrije Universiteit Amsterdam, Department of General Practice, de Boelelaan 1117, Amsterdam, the Netherlands; Amsterdam Public Health Research Institute, Health Behaviors & Chronic Diseases, Amsterdam, the Netherlands; School of Health Sciences, University of East Anglia, United Kingdom; Faculty of Medicine and Health Sciences, Keele University, Staffordshire, United Kingdom; Department of Research and Innovation, Midlands Partnership University NHS Foundation Trust, Stafford, United Kingdom; Amsterdam UMC location Vrije Universiteit Amsterdam, Department of Medical Psychology, Boelelaan 1117, Amsterdam, The Netherlands; Amsterdam UMC location Vrije Universiteit Amsterdam, Department of Clinical Pharmacology and Pharmacy, Boelelaan 1117, Amsterdam, The Netherlands; Amsterdam Public Health Research Institute, Aging & Later Life, Amsterdam, the Netherlands; Nuffield Department of Primary Care Health Sciences, University of Oxford, Oxford, United Kingdom; Amsterdam UMC, Department of Epidemiology and Data Science, de Boelelaan 1117, Amsterdam, the Netherlands

**Keywords:** Medication adherence, process evaluation, implementation, community pharmacy, type 2 diabetes mellitus

## Abstract

**Introduction:** The aim of this study was to evaluate the implementation and the potential of a pharmacist-led personalised intervention program, aimed at improving medication adherence to oral diabetes and/or hypertension medication in people with type 2 diabetes (T2D), to be normalized in daily clinical practice.

**Material and methods:** A parallel-group randomized controlled trial (RCT) was conducted in 68 participants in 26 community pharmacies from the Netherlands (NL) and the UK. This manuscript reports on a multi-method process analysis that was performed parallel to the six-month follow-up of the RCT. Quantitative data, including data from the data management platform, were analyzed with descriptive statistics. Qualitative data were collected via semi-structured interviews with purposively selected pharmacists (high/low numbers of recruited participants) and participants (high/low satisfaction) and audio-recorded consultations between pharmacists and participants. Semi-structured interviews were analyzed using the Framework approach, the Medical Research Council process evaluation framework of complex interventions and Extended Normalization Process Theory. Audio-recorded consultations were analyzed using the OPTION-12 scale and a self-composed checklist.

**Results:** The *fidelity* and *dose* of the intervention program were assessed as high. Consultations were conducted according to the study protocol. Participants’ satisfaction with the intervention modules varied, but overall, participants were satisfied with the clinical medication review, medication schedule, medication dispensing system and referral to their primary care physician. The *reach* was assessed as low, resulting from selection procedure’ difficulties and only 68 out of the intended 300 participants were recruited. Selection procedure *adaptations* did not increase the reach. The *potential* of the intervention program was assessed as high, since pharmacists valued the development of a medication adherence intervention. *Contribution* was also assessed as high, because of collective action from stakeholders. Finally, *capability* and *capacity* were assessed as moderate, due to material resource and workability barriers.

**Discussion:** In its current form the pharmacist-led personalised intervention program cannot be implemented and normalized into daily care. In this process evaluation several barriers including the selection procedure and recruitment of participants have been identified that need to be resolved before the effectiveness of this personalised intervention program can be re-evaluated. Future research should therefore focus on alternative methods for engaging people with T2D with insufficient medication adherence in medication adherence interventions.

## Introduction

Insufficient medication adherence is a common health problem in people with chronic diseases, such as type 2 diabetes (T2D) (1, 2). In the past decades numerous interventions have been developed to improve medication adherence in people with T2D (3). A systematic review of such interventions reported that those incorporating multiple strategies to address insufficient medication adherence are more effective than interventions using one strategy, but overall effects were marginal and sometimes conflicting (3). Various explanations have been suggested, such as not targeting people with insufficient medication adherence and using interventions that are not personalised (3–5).

The ImproviNg Treatment adhErence iN people with diabeteS mEllitus (INTENSE) study was a parallel-group randomized controlled trial (RCT) conducted in community pharmacies in the Netherlands (NL) and the United Kingdom (UK). The aim of the study was to improve medication adherence to oral diabetes and/or hypertension medication in people with T2D. The intervention group received a pharmacist-led personalised intervention program that could consist of (i) brief messaging, (ii) clinical medication review, (iii) addressing the medication schedule, (iv) reminding messaging, (v) medication dispensing systems, (vi) smart messaging, (vii) referral to their primary care physician and (viii) unguided web-based self-help application for low mood. The control group received usual care and access to a publicly available diabetes website. Participants were followed up for six months (6). The pharmacist-led personalised intervention program did not demonstrate a clinically significant improvement in medication adherence or the secondary outcomes such as systolic blood pressure and HbA1c level (7).

RCTs can be used to evaluate the effectiveness of interventions (8). Process evaluations alongside clinical trials afford better understanding of the fidelity and quality of the implementation, causal mechanisms and contextual factors associated with varying study outcomes (8). These insights can inform adaptations of the intervention and guide policy makers in decision-making for adoption of interventions. In this manuscript, to guide the process evaluation, we used the Framework approach, the Medical Research Council (MRC) process evaluation framework for complex interventions (8) and Extended Normalization Process Theory (ENPT) (9).

The aim of this study was to evaluate the implementation and the potential of a pharmacist-led personalised intervention program, aimed at improving medication adherence to oral diabetes and/or hypertension medication, in people with T2D to be normalized in daily clinical practice.

## Methods

The research protocol was approved by the Medial Ethical Committee of the VU Medical Center Amsterdam (reference 2018.160) and the Health Research Authority and Health and Care Research Wales (reference 19/WA/0275). Written informed consent was obtained from participants.

### Study design

The design of the INTENSE study has been described in more detail in the study protocol (6). This process evaluation was conducted parallel to the RCT. Data were obtained during the six-months trial period and after trial completion.

### Conceptual frameworks

The MRC process evaluation framework for complex interventions guided this process evaluation with ENPT providing the underlying theory. *Implementation* was assessed in terms of (i) *fidelity*, i.e. whether the intervention was delivered as intended, (ii) *dose*, i.e. what was the quantity of the intervention and (iii) *reach*, i.e. did the intended population come into contact with the intervention and how. In addition, we assessed if any *adaptations* were made to the pharmacist-led personalised intervention program and evaluated whether *contextual factors* affected implementation. *Mechanisms of impact*, *description of the intervention and its causal assumptions* and *outcomes* were not assessed in this process evaluation (8).

In addition, the constructs of ENPT were used to structure qualitative data. ENPT focusses on the effort that stakeholders put into an intervention in order for a complex intervention to normalize in daily clinical practice and consists of four core elements, namely *potential*, *capacity*, *capability* and *contribution* (9). The first three elements are *potential*. i.e. the social-cognitive resources that are available to pharmacists, *capability*, i.e. possibilities that are offered by the intervention and *capacity*, i.e. the social-structural resources available to pharmacists. Together these three elements form *contribution*, which indicates what pharmacists do to implement the intervention (9). Research questions were developed and were structured around the MRC process evaluation framework of complex interventions and ENPT (**Table 1)**.

**Table 1.** Overview of the conceptual framework constructs, research questions and data sources. I iterviews; II, audio-recorded consultations; III, data management platform; IV, satisfaction phone call; V, administration research team

| Framework | Construct | Research question | Data source(s) | Outcomes |
| --- | --- | --- | --- | --- |
| **MRC process evaluation framework of complex interventions** | Fidelity | - Pharmacist-led personalised intervention program delivered as intended<br>- Pharmacist-led personalised intervention program that was delivered | I, II, III, IV, V | Shared decision-making was conducted, although sometimes to a limited extent.<br>The datamanagement platform was used to guide the consultations and was a facilitator for conducting the consultations according to the study protocol.<br>Fidelity barriers for participants were that consultations were sometimes short and lacked depth and for pharmacists that the consultation did not fit their own consultation style and sometimes help was needed with the selection procedure. |
|  | Dose | - Pharmacist-led personalised intervention program that was delivered<br>- Pharmacist-led personalised intervention program that was received<br>- Participants' engagement with the pharmacist-led personalised intervention program | I, II, III, IV, V | The clinical medication review, brief messaging and Unguided web-based Self Help Application for low mood were offered and selected the most. Participants' satisfaction with the clinical medication review, medication schedule, medication dispensing system or referral to their primary care physician were highest. For the brief messaging, smart messaging and Unguided web-based Self Help Application for low mood satisfaction differed or was low. |
|  | Reach | - Recruitment of the intended study population | I, III, V | Very few participants agreed to participate in the study. Pharmacists experienced difficulties with the selection procedure, which was time-consuming and did not select the wright patient sample. Participants often did not found themselves in need of the intervention. This was confirmed by pharmacists that mentioned that participants often experienced few problems with their medication intake. |
|  | Context | - Reasons for participation |  | Contextual facilitators were that the health care professional send out the |
|  |  | - Barriers for participation |  | invitations and barriers were that the information consent procedure was on paper and impersonal. |
|  | Adaptations | - Adaptations that were made | V | Multiple adaptations were made to the selection procedure, i.e. refinement of the search strategy and adjustment of the inclusion and exclusion criteria, but this did not improve participant recruitment. |
| **ENPT** | Potential | - Commitment with the pharmacist-led personalised intervention program<br>- Assessment of change efficacy of the pharmacist-led personalised intervention program | I | Pharmacists overall valued that an intervention program was developed to support people with T2D with their medication adherence and it helped them in offering better health care. Team members were committed to the study. |
|  | Capacity | - Changes needed in working processes<br>- Feasibility of the changes needed in the working processes of the pharmacy | I | Pharmacists valued the intervention materials and pharmacist-led personalised intervention program. However, there were multiple material resource barriers, including the patient selection procedure that was extensive and missed eligible patients in the UK. In addition, the training was time-consuming, there was limited time and trained-staff available in the pharmacy, and in the NL financial reimbursement was insufficient. Other material resource barriers were that the informed consent form was too difficult and extensive for the overall target population. In general, pharmacists saw it as their role to support patients with medication adherence, but also mention that this is a shared responsibility with the patient and other health care professionals. Cognitive resources were sufficient, with an exception of knowledge on how to offer the mood intervention. |
|  | Capability | - Experiences with the pharmacist-led personalised intervention program<br>- Workability of the pharmacist-led personalised intervention program | I | The pharmacy consultation was deemed workable in terms of time investment and during the COVID-19 pandemic. However, in the NL, time investment of the patient selection was high and overall the study was labor-intensive for the limited number of participants. In the UK, the task distribution of the different stakeholders was a barrier in the workability. Opinions on whether the intervention program could be integrated into clinical practice were conflicting. |
|  | Contribution | - Distribution of tasks related to the study | I | There was an understanding of the pharmacist-led personalised intervention program, indicating good coherence. Conversely, pharmacists expressed barriers for coherence with regard to the selection procedure and wondered why this was not performed differently. Collective action of the different stakeholders was good. |

### Data collection

Multiple data sources, consisting of both quantitative and qualitative data, were collected in the NL and the UK (**Table 1**).

First, semi-structured interviews with a purposive sample of pharmacists and participants were conducted, either face-to-face or via telephone. The purposive sample consisted of pharmacists with high and low numbers of recruited participants and participants with high and low satisfaction with the interventions. Semi-structured interviews were conducted by research team members and were guided by topic lists informed by the conceptual frameworks.

Second, we used a random sample of intervention pharmacy consultations in the intervention group, which were audio-recorded by the pharmacist. During the consultation questions from the Adapted Quick Barriers Scan (QBS) questionnaire, i.e. questionnaire aimed at identifying barriers for medication adherence, and the intervention modules were discussed.

Third, the data management platform was used by researchers and the pharmacy team to record participants’ data in the intervention and control group. This included participants’ non-adherence profiles and selected intervention modules.

Fourth, approximately one month after the start of the pharmacist-led personalised intervention program, a researcher (NL) or English pharmacist (UK) called the participants from the intervention group to conduct a satisfaction phone call. In this call participants were asked, according to a telephone protocol, if the intervention modules were initiated, reason(s) if not initiated, satisfaction with the intervention modules, reason(s) for satisfaction, and if changes were needed to be made to the intervention modules.

Fifth, we recorded participants’ and pharmacists’ descriptive data, logistics and flow of participants throughout the study and whether changes were made to the study protocol.

### Data analyses

Quantitative data were analyzed using descriptive statistics. Continuous variables were presented as means with standard deviations and categorical variables as frequencies with percentages. When data were missing they were excluded from the analyses. Analyses were conducted in SPSS Statistics 26.

Both the semi-structured interviews and audio-recorded pharmacy consultations were transcribed verbatim. Semi-structured interviews were analyzed using the *implementation* and *context* dimensions from MRC process evaluation framework of complex interventions and all core elements from the ENPT. First, all transcripts were openly coded by two independent researchers, to see whether the data would fit the constructs from both frameworks. Second, codes were arranged to themes by using the constructs of the MRC process evaluation framework of complex interventions for participant and pharmacist interviews and the ENPT for interviews with pharmacists. Arrangement of codes to themes was performed by two researchers (MLG and PJME), using MAXQDA (10).

For the analysis of the audio-recorded consultations, transcripts were scored on the use of shared-decision making with the OPTION-12 scale (**Table 3**), a tool to assess the level of shared-decision making of audio-recorded consultations in general practices. The scale consists of 12 items that are scored on a 5-point scale, ranging from ‘not observed behaviour’ to ‘behaviour that is observed and executed to a high standard’, and has an intraclass correlation coefficient of 0.77 (11). In addition, a non-validated checklist was developed to assess whether consultations were performed according to study protocol. The checklist comprised of five items (**Table 4**), including whether the pharmacist used the data management platform during the consultation. Items were scored on a dichotomous scale, i.e. yes or no, by two independent researchers (MLG and a pharmacy master student).

Quantitative results were expressed numerically and qualitative results were presented with descriptive statistics.

## Results

In the INTENSE study, 26 community pharmacies participated (23 NL and 3 UK), including a total of 68 participants (47 NL and 21 UK). Participant recruitment was undertaken for the planned eight months in the UK and 16 months in the NL. **Figure 1** provides the flowchart of study participants in the NL and the UK. Both the intervention and control group consisted of 34 participants. For this process evaluation, data management platform data, satisfaction phone call, and administration research team were analyzed for the intervention group. The patients in the intervention group consisted of 26 men (76.5%), with a median age of 66.5 years [59.8 to 73.0]. In addition, we analyzed semi-structured interviews of 9 pharmacists (5 NL and 4 UK) and 9 participants (5 NL and 4 UK) and 6 audio-recorded consultations (4 NL and 2 UK). The pharmacist sample that participated in the semi-structured interviews consisted of 1 man (11%) and the participant sample consisted of 8 men (89%).

**Figure 1.**
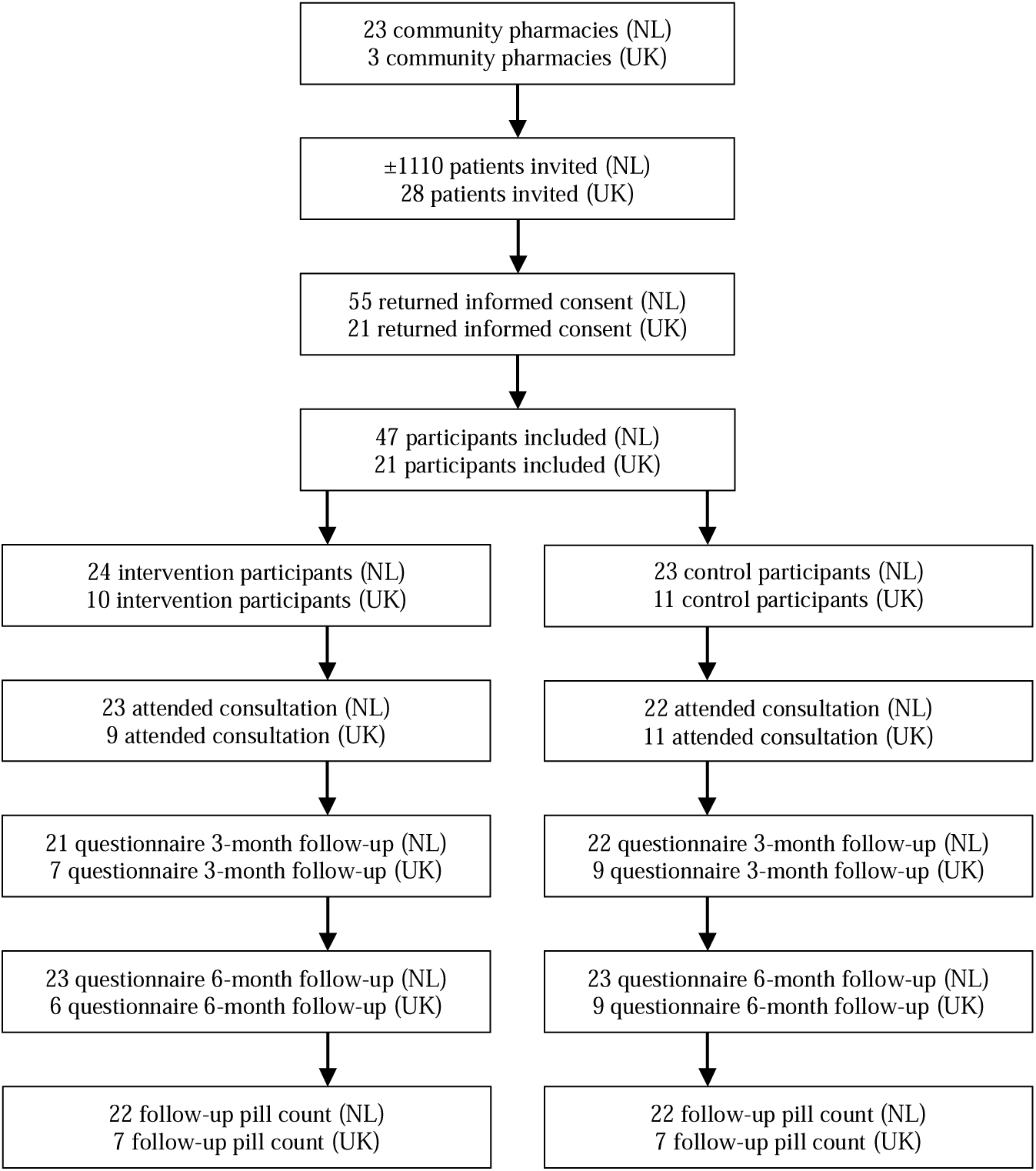
The flowchart of the INTENSE study participants for the Netherlands and the UK.

### Implementation and context according to the MRC framework

#### Fidelity

##### Semi-structured interviews

For the delivery of the intervention according to the study protocol, i.e. the fidelity, some participants indicated that pharmacists did more than solely offer the interventions.

Participants for instance pointed out that pharmacists sometimes provided more general information on the working mechanisms of medication (**Table 2, quote 1**). Most participants mentioned that they were satisfied with the pharmacist’ consultation and the personal contact with the pharmacist was seen as a positive aspect of the pharmacist-led personalised intervention program. In contrast, several participants stated as a barrier for the delivery of the intervention that they felt like the pharmacist was not completely committed to the study, because they went through the consultation quickly, did not explain the intervention modules in detail and the consultation lacked depth (**Table 2, quote 2**). In a single case, the consultation appointment was cancelled several times by the pharmacist.

**Table 2.** Quotes from the semi-structured interviews. P, participant; PH, pharmacist; NL, the Netherlands; UK, United Kingdom

| Number | Quote |
| --- | --- |
| 1. | “She also indicated that if I skipped once what the effect of the medicine would be. She actually just indicated how the medicine worked.” – P1 NL |
| 2. | “The consultation was fine. I did not feel like a whole lot came out of it so to speak, that a lot of improvement would come out of it.” – P4 NL |
| 3. | “My biggest fear is that you will get to a point where certain medicines no longer work properly or that there will be side effects.” – P4 NL |
| 4. | “It wasn’t very helpful for me. Everything that was mentioned I already do.” – P7 UK |
| 5. | “We felt that we were missing patients. Something wasn’t right.” – PH9 UK |
| 6. | “Well, I might as well help if it helps people to understand more about medications, diabetes, high blood pressure. Let’s give it a go.” – P6 UK |
| 7. | “Perhaps you could shift more towards digital versions, but perhaps that might be more difficult for the target audience, that could be.” – P2 NL |
| 8. | “Actually spoke to you when you visit them, that would help as well. A more personal approach then just a letter. Some people might put the letter down and then forget about it and not bother anymore.” – P8 UK |
| 9. | “I think it’s pharmacists are really well placed to help people with compliance issues, or anti diabetic medication.” – PH9 UK |
| 10. | “And sometimes you have people who you select and you think well no I should not send them the letter, because you know they will not understand.” – PH5 NL |
| 11. | “I think it’s a shared responsibility between the patient and their carers. The prescriber and the pharmacist, and the fact that it’s a shared responsibility, kind of, everyone can do it.” – PH9 UK |
| 12. | “The guided self-help application I didn’t see you know it might be useful to actually see what the modules entail just to so you can explain it to the patients a little bit better.” – PH7 UK |
| 13. | “The conversation is much more efficient, because you can tackle things faster. In terms of time-investment I thought it was okay, it actually made the conversation go much smoother.” – PH1 NL |
| 14. | The sheet (for checking the inclusion and exclusion criteria) was a bit too extensive to complete.” – PH4 NL |
| 15. | “I’m not really sure what impact brief messages that’s great on a day-to-day basis but is that something that can continue long-term I’m not sure.” – PH7 UK |
| 16. | “I felt that there wasn’t enough follow-up with surgery on making appointments for patients, because at the follow-up consultation when people should have had a medication review that did happen at that point. That was a bit disappointing.” – PH7 UK |

**Table 3.**
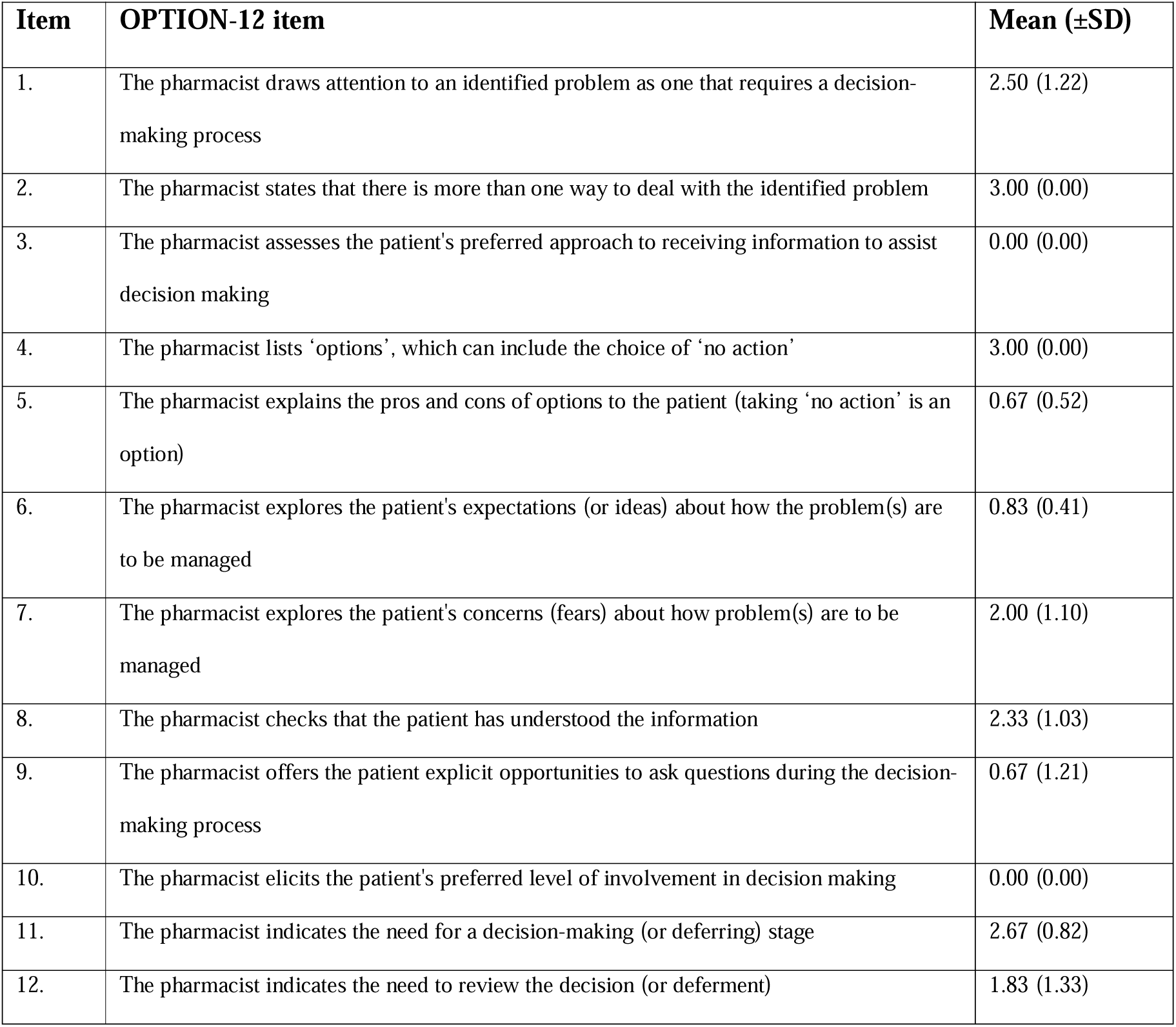
Observed mean numbers and standard deviations per OPTION-12 item, with ‘clinician’ from the original OPTION-12 scale replaced by ‘pharmacist’.

| Item | OPTION-12 item | Mean (±SD) |
| --- | --- | --- |
| 1. | The pharmacist draws attention to an identified problem as one that requires a decision-making process | 2.50 (1.22) |
| 2. | The pharmacist states that there is more than one way to deal with the identified problem | 3.00 (0.00) |
| 3. | The pharmacist assesses the patient's preferred approach to receiving information to assist decision making | 0.00 (0.00) |
| 4. | The pharmacist lists 'options', which can include the choice of 'no action' | 3.00 (0.00) |
| 5. | The pharmacist explains the pros and cons of options to the patient (taking 'no action' is an option) | 0.67 (0.52) |
| 6. | The pharmacist explores the patient's expectations (or ideas) about how the problem(s) are to be managed | 0.83 (0.41) |
| 7. | The pharmacist explores the patient's concerns (fears) about how problem(s) are to be managed | 2.00 (1.10) |
| 8. | The pharmacist checks that the patient has understood the information | 2.33 (1.03) |
| 9. | The pharmacist offers the patient explicit opportunities to ask questions during the decision-making process | 0.67 (1.21) |
| 10. | The pharmacist elicits the patient's preferred level of involvement in decision making | 0.00 (0.00) |
| 11. | The pharmacist indicates the need for a decision-making (or deferring) stage | 2.67 (0.82) |
| 12. | The pharmacist indicates the need to review the decision (or deferment) | 1.83 (1.33) |

**Table 4.** Observed numbers and percentages per item of self-composed INTENSE item list.

| Item | INTENSE scoring items | N (%) |
| --- | --- | --- |
| 1. | The pharmacist uses the data management platform during the consultation. | 6 (100%) |
| 2. | The pharmacist discusses the purpose of the consultation with the patient. | 3 (50%) |
| 3. | The pharmacist discusses the answers of the Adapted QBS questionnaire with the patient. | 5 (83%) |
| 4. | The pharmacist discusses the interventions the patients qualifies for. | 6 (100%) |
| 5. | The pharmacist discusses with the patient which interventions the patient wants to use. | 6 (100%) |

One fidelity barrier was identified in the interviews with pharmacists, namely that assistance from the pharmacy team was needed to carry out the patient selection procedure.

##### Audio-recorded consultations

Consultations were scored on the level of shared-decision making with the OPTION-12 scale (**Table 3)**. Two items, namely “The pharmacist states that there is more than one way to deal with the identified problem” and “The pharmacist lists options, which can include the choice of no action” were reported as reaching a good standard in all six consultations. The items “The pharmacist assesses the patient’s preferred approach to receiving information to assist decision making” and “The pharmacist elicits the patient’s preferred level of involvement in decision making” were not observed in any consultation. In addition, consultations were scored with the self-composed checklist (**Table 4)**. In the data-management platform the assigned non-adherence profiles and corresponding intervention modules were accessible for pharmacists. In all audio-recorded pharmacy consultations the online data-management platform was used as a guidance. Also, we observed that the pharmacist mentioned the eligible intervention modules and that they discussed with the participant which interventions would be initiated. In five out of six consultations, the pharmacist discussed the Adapted QBS questionnaire and in only three out of six consultations the purpose of the consultation was mentioned.

##### Data management platform

Of the 34 intervention participants, 15 participants were assigned to the medication non-adherence profile *Knowledge and perceptions*, 20 participants to *Practical problems*, 16 participants to *Side effects*, 25 participants to *Negative mood and beliefs*. Five participants did not have a non-adherence profile. Of these participants, 15 chose *brief messaging*, 12 *clinical medication review*, 4 *medication schedule*, 5 *reminder messaging*, 2 *medication dispensing systems*, 2 *smart messaging*, 1 *referral to their primary care physician*, and 14 *unguided web-based self-help application for low mood* (**Table 5**).

**Table 5.** Offered and selected intervention modules and participants’ satisfaction. * 7.3% of the satisfaction data was missing and excluded from analysis

| **Intervention module** | **Times offered** | **Times selected** | **Satisfaction\*** |
| --- | --- | --- | --- |
| Brief messaging | 28 (20 NL / 8 UK) | 15 (11 NL / 4 UK) | Very dissatisfied: 2<br>Dissatisfied: 1<br>No clear opinion: 4<br>Satisfied: 4<br>Very satisfied: 1<br>Not initiated: 3 |
| Clinical medication review | 31 (22 NL / 9 UK) | 12 (10 NL / 2 UK) | Very dissatisfied: 0<br>Dissatisfied: 0<br>No clear opinion: 0<br>Satisfied: 6<br>Very satisfied: 4<br>Missing: 2 |
| Medication schedule | 20 (15 NL / 5 UK) | 4 (4 NL / 0 UK) | Very dissatisfied: 0<br>Dissatisfied: 0<br>No clear opinion: 0<br>Satisfied: 3<br>Very satisfied: 0<br>Not initiated: 1 |
| Reminding messaging | 19 (14 NL / 5 UK) | 5 (5 NL / 0 UK) | Very dissatisfied: 1<br>Dissatisfied: 0<br>No clear opinion: 0<br>Satisfied: 3<br>Very satisfied: 1 |
| Medication dispensing systems | 20 (15 NL / 5 UK) | 2 (2 NL / 0 UK) | Very dissatisfied: 0<br>Dissatisfied: 0<br>No clear opinion: 0<br>Satisfied: 2<br>Very satisfied: 0 |
| Smart messaging | 20 (15 NL / 5 UK) | 2 (1 NL / 1 UK) | Not initiated: 2 |
| Referral to their primary care physician | 17 (14 NL / 3 UK) | 1 (1 NL / 0 UK) | Very dissatisfied: 0 |
|  |  |  | Dissatisfied: 0<br>No clear opinion: 0<br>Satisfied: 1<br>Very satisfied: 0 |
| Unguided web-based Self Help Application for low mood | 27 (19 NL / 8 UK) | 14 (7 NL / 7 UK) | Very dissatisfied: 2<br>Dissatisfied: 1<br>No clear opinion: 1<br>Satisfied: 1<br>Very satisfied: 0<br>Not initiated: 7<br>Missing: 2 |

#### Dose

##### Semi-structured interviews

No information was provided on dose.

##### Satisfaction phone call

Table 5 provides an overview of the participants’ satisfaction with the intervention modules. All participants who received a clinical medication review, medication schedule, medication dispensing system or referral to their primary care physician were either satisfied or very satisfied with the intervention module. Participants were satisfied with the clinical medication review because of the openness and attentiveness during the consultation, the additional information received about medicines and follow-up initiation. Satisfaction with the medication schedule and medication dispensing systems were caused by the fact that these interventions were indeed provided. Satisfaction with the referral to their primary care physician was caused by smooth referral and a good consultation with the GP.

Four out of five participants who received reminding messaging were satisfied or very satisfied, because it served as a reminder, especially for new medication. However, one participant indicated to be very dissatisfied due to technical difficulties with the initiation of the module. Several participants mentioned that they would have preferred continuation of reminders after study completion (**Table 5**).

For the brief messaging intervention module satisfaction differed. Five out of fifteen participants scored the intervention modules with satisfied or very satisfied. Reasons for satisfaction were that the messages kept them sharp and also served as a form of reminder. Three out of fifteen participants scored the intervention module with dissatisfied or very dissatisfied, because of superficial content and no improvement in participants’ knowledge.

The smart messaging intervention module was selected by two participants. Both times the intervention module was not initiated, due to logistic problems or the device not able to fit all the participants’ medicines.

Finally, fourteen participants selected the unguided web-based self-help application for low mood, of which seven never started the program. One participant was satisfied with the program, because of its informative character. Three out of seven participants that started using the application were dissatisfied or very dissatisfied. Explanations that were given included that participants felt like they were not getting anything out of it or that the application gave them negative or somber feelings. Two participants had missing data on their satisfaction with the unguided web-based self-help application for low mood (**Table 5**).

#### Reach

##### Semi-structured interviews

During the interviews participants were not asked whether they deemed themselves to have insufficient medication adherence. Overall, participants often indicated to experience few problems with their medication intake and as such little improvement in medication adherence could be achieved. Four participants however mentioned that they sometimes forget to take their medicines, for instance in the evening. One participant told “sometimes I have too much medicine in stock” and a second participant expressed the “fear of having side effects and that the medicines might not work properly anymore in the future” (**Table 2, quote 3**). In contrast, many participants indicated that they never forget to take their medicines and that the information they received by means of the pharmacist-led personalised intervention program was already known to them (**Table 2, quote 4**). One participant however did mention that after the pharmacy consultation, he started using the reminder text messages and benefited from it in terms of a more regular intake.

Almost all pharmacists mentioned the patient selection procedure as a major barrier in the study and their dissatisfaction with the number of participants recruited. Pharmacists stated that very few patients were eligible for recruitment, possibly due to the exclusion of patients using medication-intake supporting devices provided by the pharmacy. Primarily in the UK, pharmacists indicated that they felt that the selection procedure did not lead to selection of the appropriate patients (**Table 2, quote 5**) and one UK pharmacist even withdrew because of this. Finally, participants that were recruited often indicated to experience few problems with their medication intake and as such little improvement in medication adherence could be achieved.

##### Administration research team

Approximately 1110 patients were invited to participate in the NL of whom 47 consented. In the UK, 28 patients were invited to participate of whom 21 consented. This provided a total of 68 participants. Patient barriers for participation were unclear since the Medical Ethical Committees restricted us to from obtaining reasons for non-participation in the patients who did not return an informed consent form.

#### Adaptations

##### Administration research team

In the NL, inclusion and exclusion criteria were altered, allowing people with a dispensing score of 90% or less to participate instead of the initial 80% or less. Also, pharmacies were allowed to recruit people if at least one note about insufficient medication adherence was made in the pharmacy system in the last year, even if dispensing data were in order.

Additionally, two exclusion criteria were dropped, allowing people with insulin or repeat dispensing service from the pharmacy to participate. These adaptations did not substantially increase participant recruitment.

#### Context

##### Semi-structured interviews

Most participants indicated that they did not participate for personal benefit. The main motivator seemed to aid research in general as well as other people with T2D (**Table 2, quote 6**). Participants pointed out that they were more willing to participate, because they were invited by their health care provider. The informed consent procedure was mentioned as a barrier to participate. One participant indicated that the formal informed consent procedure could have been easier when conducted digitally instead or via post (**Table 2, quote 7**).

Another participant deemed the patient information sheet and informed consent procedure impersonal and would have preferred a verbal procedure by their health care provider (**Table 2, quote 8**).

### Normalization according to the ENPT

#### Potential – Individual intentions

##### Semi-structured interviews pharmacists

Pharmacists provided several reasons for study participation. First, insufficient medication adherence is a major issue in clinical practice, which needs to be addressed. Pharmacies often indicated that they want to offer more interventions to their patients with insufficient medication adherence (**Table 2, quote 9**) and this study provided them with a possibility to do so. In addition, multiple pharmacists indicated that they participated in the INTENSE study to offer better health care, showing the patient what the pharmacist can do, and as such build relationships with patients, researchers and other health care professionals. In contrast, the complexity of medication non-adherence was also indicated as a barrier for participation.

#### Potential – Collective commitment

##### Semi-structured interviews pharmacists

Several pharmacists said that there was collective commitment for participation in the study and that pharmacy staff was positive about participation. In the UK, one pharmacist mentioned insufficient collective commitment by their participating primary care physician.

#### Capacity – Material resources

##### Semi-structured interviews pharmacists

Overall, pharmacists were positive about the intervention materials. The protocol was easy to use, the training was clear and the data-management platform was easy to use during the consultation. The pharmacist-led personalised intervention program was perceived as positive since it added on current interventions for insufficient medication adherence.

In contrast, there were also negative comments about the materials. Some pharmacists found the training videos to be too long. In the UK, pharmacists additionally received an obligatory Good Clinical Practice (GCP) training, which also contained irrelevant information for this current project. Additionally, the patient selection procedure was perceived as too extensive, and especially in the UK doubts were expressed on whether the right patients were selected with the selection algorithm in medical records of primary care physicians. Other barriers for conducting the intervention program were limited time in the pharmacy and insufficient numbers of trained staff. Pharmacists in the NL indicated that if this intervention were integrated into standard care, additional financial reimbursement would be needed. Finally, multiple pharmacists in the NL thought that the patient information letter and informed consent form were too difficult and extensive for the target population (**Table 2, quote 10**).

#### Capacity – Social roles

##### Semi-structured interviews pharmacists

Pharmacists indicated that supporting patients with medication adherence fits their social role. They specified a signaling role, because they have insights into patients’ adherence. It was mentioned several times that providing information to patients during the first and second dispatch visit, supports medication adherence. However, it was also mentioned that adherence support should be a shared responsibility of the patient, pharmacist and medical doctor and in the NL the nurse practitioner (**Table 2, quote 11**).

#### Capacity – Social norms

##### Semi-structured interviews pharmacists

No information was provided on social norms.

#### Capacity – Cognitive resources

##### Semi-structured interviews pharmacists

Despite the complexity of medication non-adherence, pharmacists indicated having sufficient knowledge on medication adherence and how to address it in the pharmacy. Conversely, some pharmacists reported a lack of knowledge for offering a mood intervention (**Table 2, quote 12**).

#### Capability – Workability

##### Semi-structured interviews pharmacists

The pharmacy consultation was deemed workable in terms of time investment. Questions from the Adapted QBS were efficiently discussed, because the questionnaire was filled out beforehand by participants. As such, the pharmacist was able to prepare the consultation and did not need to discuss irrelevant questions from this questionnaire (**Table 2, quote 13**). In addition, the consultation was workable during the COVID-19 pandemic and digital components fit current times.

In the NL, the patient selection was indicated as unworkable. For each potential participant the pharmacist needed to check inclusion and exclusion criteria and this was time-consuming (**Table 2, quote 14**). Besides, many pharmacies were only able to recruit very few participants. The amount of activities that needed to be performed for this limited number of participants was high and as such the study was labor-intensive. It was also mentioned by the pharmacists that the informed consent procedure hindered the intervention.

In the UK, the distribution of tasks related to the intervention between the research team, pharmacy and primary care physicians was sometimes unclear to the pharmacist. This could have been resolved by having a meeting with all stakeholders at the start of the study. One pharmacist expressed that the study did not feel workable, because they felt it was not yet thought out. When the pharmacist had their own ideas on how to improve participants recruitment they were restricted by research regulations, which made it more difficult to make these changes in practice. An example being the long and impersonal patient information sheet, dictated by the ethical committees.

#### Capability – Integration

##### Semi-structured interviews pharmacists

Opinions on whether the pharmacist-led personalised intervention program could be integrated into daily clinical practice were conflicting. Some pharmacists indicated that the intervention could be scaled, but some stated it cannot fit into routine care. One pharmacist specifically indicated that the questions from the Adapted QBS gave more depth to the consultations, which would be an asset to clinical practice. Another pharmacist indicated that the consultation set-up did not fit the pharmacists’ own consultation style

#### Contribution – Coherence

##### Semi-structured interviews pharmacists

For the intervention modules, one pharmacist expressed a coherence barrier, namely that they did not see how brief messages could be effective in improving medication adherence on the long-term (**Table 2, quote 15**). On the other end, one pharmacist expressed a clear understanding of the underlying working mechanisms of the mood intervention. For the patient selection, pharmacists mostly expressed coherence barriers. These barriers often focused on the selection of participants and why this was not conducted differently. In addition, the limited number of participants raised concerns with the pharmacists.

#### Contribution – Cognitive participation

##### Semi-structured interviews pharmacists

No information was provided on contribution.

#### Contribution – Collective action

##### Semi-structured interviews pharmacists

For collective action, several views were expressed. Overall, pharmacists mentioned that they had good collaboration with the research team and team of primary care physicians, because these stakeholders were involved and communicated well. Negatively, one pharmacist indicated that collective action was limited for the team of primary care physicians, since they did not schedule a clinical medication review, hampering the pharmacist in performing the satisfaction phone call (**Table 2, quote 16**).

#### Contribution – Reflexive monitoring

##### Semi-structured interviews pharmacists

No information was provided on contribution.

## Discussion

In the INTENSE study, the pharmacist-led personalised intervention program did not improve medication adherence in people with T2D (7). In this process evaluation we assessed the implementation of this intervention program and showed that most of the intervention components were implemented as intended. However, limited reach hampered the effectiveness analysis due to insufficient statistical power. Overall, the process analysis shows that in its current form the pharmacist-led personalised intervention program cannot be implemented in daily clinical practice.

### Reach

In our analysis, several factors were identified that led to limited reach. First, the selection procedure of participants via pharmacy records or medical records on refill data (6) was insufficient, as we encountered difficulties with the refinement of the search strategy in the UK and as a result not all eligible patients were selected. This is in line with a recent study from Hogervorst et al., aimed at improving medication adherence in people with T2D in the NL, in which health care providers also mentioned that pharmacy refill data was not reliable for identifying insufficient medication adherence, because repeat dispensing distorts pharmacy refill data (12).

Second, pharmacists questioned participants’ eligibility. In the Cardiovascular medication non-Adherence Tailored Intervention CATI study (5), a precursor for this study, pharmacists also mentioned this as a barrier (13). Therefore, we selected more rigid inclusion and exclusion criteria. Interestingly, this did not lead to an improvement in the eligibility of participants. Participants often felt that they were not in need of the intervention program, which was supported by the interviews with pharmacists. This is potentially caused by the fact that people that are willing to participate in clinical research are not the ‘typical’ non-adherent patient. Selection bias might be present, as participating in this study involved a lot of steps, including the informed consent procedure and multiple questionnaires. However, since we were restricted from obtaining reasons for non-participation we do not know whether this procedure and questionnaires hampered the inclusion. Other reasons, previously identified in research included patients’ concerns that their treatment regimen will be altered or that medication will be taken away (14, 15).

Third, pharmacists indicated that consultations were workable during the COVID-19 pandemic, potentially because the research team was able to adapt the study set-up. However, we do hypothesize that the pandemic substantially impacted recruitment. Many treatments for chronic conditions were interrupted or affected during the pandemic due to reduced adherence rates (16). This implies that there could have been a larger group of people eligible for the study. However, this did not increase participant numbers, possibly due to people being reluctant to visit their health care provider out of fear of a COVID-19 infection (17). However, as we do not know the reasons for non-participation, we cannot rule out any selection bias.

Fourth, both participants and pharmacists pointed out that the informed consent procedure was a barrier for participation. Pharmacists described the patient information letter and informed consent form to be too difficult and extensive. The information letter and informed consent form were developed based on legally required templates. The informed consent procedure is a mandatory step in research, we agree that the currently required regulatory templates are difficult, especially for patients with a language barrier or low educational level, supported by several other studies (14, 18). We hypothesize that in our study, the required informed consent procedures led to reduced participation rates and selection bias.

### Pharmacist-led personalised intervention program

We also found positive factors for implementing a pharmacist-led personalised intervention program in daily clinical practice. First, pharmacists were willing to use the program, in order to deliver high quality care and support medication adherence, and participants valued the personal contact with their pharmacist. This personal approach was also mentioned as a facilitator in the recruitment process, which is in line with other studies (14, 18, 19). In addition, pharmacists indicated the training and intervention materials as facilitators for the delivery of the pharmacist-led personalised intervention program, also in line with previous studies (13, 20). Moreover, participants’ satisfaction with the intervention modules differed, but participants were satisfied with the clinical medication review, medication schedule, medication dispensing system and referral to their primary care physician. Interestingly these are all intervention modules initiated or carried out by either the pharmacist or primary care physician. Finally, pharmacists in our study indicated that adherence support should be a shared responsibility of the patient, pharmacy and medical doctor, supported by previous studies (21, 22). Future studies should therefore also use a multidisciplinary approach.

In contrast, we also identified additional barriers for the delivery of the intervention program. First, patients did not consider all intervention modules helpful, for instance the information provided by the brief messaging intervention was already known to them. Second, pharmacists should have sufficient time for the intervention (20), however the time invested in the current set-up was too high, especially for the limited amount of participants. Third, there must be adequate financial compensation for carrying out the intervention. Finally, collaboration with the UK primary care physician teams was insufficient, which is often seen as a barrier for implementation, especially for digital interventions (12).

### Limitations and strengths

This process evaluation consists of several strengths and limitations. The first limitation is that the semi-structured interviews with pharmacists did not elicit information on all ENPT constructs, although the topic lists were developed and structured around the frameworks. However, we assume that all of the relevant facilitators and barriers for implementation and normalization were mentioned and data saturation was reached as constructs were missing due to them not being relevant. Second, we used some subjective tools, for example the OPTION-12 scale to rate the level of shared-decision making in the consultations and the non-validated checklist to assess whether consultations were performed according to study protocol. It should however be noted that an objective rating of the audio-recorded consultations is unfeasible. The subjective ratings were conducted by two independent researchers, and discussed afterwards, to increase the rigor of the reported results.

The first strength is that we were able to include a random sample of audio-recorded pharmacist consultations, to provided us with insights on whether the intervention was carried out according to the study protocol, which can be a ‘black-box’ in other studies (13). A second strength is that we conducted both interviews with pharmacists and participants, which provided us with an understanding of how both stakeholders perceived the intervention program. A third strength is that we structured this process evaluation around two frameworks. The use of a framework is recommended when conducting process evaluations and by using the MRC process evaluation framework of complex interventions and ENPT we were able to analyze both the implementation of the pharmacist-led personalised intervention program and the potential of the intervention to be normalized in daily care.

### Implications for daily practice

In its current form the INTENSE pharmacist-led personalised intervention program cannot be implemented and normalized into daily care. In this process evaluation several barriers have been identified that need to be resolved, before the effectiveness of this pharmacist-led personalised intervention program can be re-evaluated. Improvement of the recruitment process and reach of health care interventions is important (18, 19). Potentially an alternative way to identify and recruit people with T2D that have insufficient medication adherence could be by purposive sampling. Purposive sampling is a method that is often applied in qualitative research and that is based on intentionally selecting patients with specific characteristics (23). This would mean that pharmacists can offer the pharmacist-led personalised intervention program to patients that they suspect to have insufficient medication adherence. However, in the NL the addition of purposive sampling did not increase participation rates. We hypothesize that another recruitment method could be to offer the intervention program to patients that indicate that they want to improve their medication intake. In addition, the personal contact with the pharmacist was mentioned as a facilitator in the recruitment process. Therefore, we suggest that the pharmacist should first provide verbal information about the intervention program. The subsequently offered patient information letter and informed consent form should be written in such a way that it is short and understandable for people with a language barrier and perhaps make use of icons or images.

Finally, the trial should be embedded into routine clinical pathways, with the trial being almost inseparable from routine care.

## Conclusion

In conclusion, the process evaluation of the INTENSE study showed that most of the intervention components were implemented according to the study protocol. However, the limited reach of the pharmacist-led personalised intervention program, due to difficulties with the selection procedure and recruitment of participants appeared a major barrier. Therefore, in its current form, the pharmacist-led personalised intervention program cannot be implemented in daily clinical practice. Future research should focus on alternative methods for the recruitment of people with T2D that have insufficient medication adherence in medication adherence studies.

## Declarations

### Data availability

Participant data cannot be supplied because no informed consent was obtained for data sharing.

### Authors’ contributions

AF, JH and PE developed the basis for the study design and protocol. FR, MdW and AAH were involved in the study design. AF, DB, HA, JH, ML-G and PE were members of the trial managing committee and met on average once every 2 months. ML-G coordinated data collection in the NL and conducted the data analyses and reporting of the study results. DB coordinated data collection and qualitative data analysis in the UK. FR, ML-G and PE drafted this manuscript. All authors read and approved the final manuscript.

## Acknowledgments

The authors would like to thank research assistants and students that provided data collection and data-analyses support.

